# The cross-sectional correlation between the oxidative balance score and cardiometabolic risk factors and its potential correlation with longitudinal mortality in patients with cardiometabolic risk factors

**DOI:** 10.1101/2023.11.10.23298401

**Authors:** Qiancheng Lai, Liu Ye, Jun Luo, Cheng Zhang, Yue Shao

**Author notes:** Correspondence author: Yue Shao, Department of Cardiothoracic Surgery, The First Affiliated Hospital of Chongqing Medical University, NO. 1 Youyi Road, Yuzhong District, Chongqing, China.

## Abstract

**Background:** This study sought to analyze the correlation between oxidative balance score (OBS) and cardiometabolic risk factors (CMRFs) and mortality in individuals with CMRFs.

**Methods:** Data were chosen from the National Health and Nutrition Examination Survey. The survey-weighted multivariable logistic regression models were implemented to explore the relationship between OBS and the risk of CMRFs. Then, Cox proportional hazard models were employed to estimate the impact of OBS on mortality in individuals with CMRFs.

**Results:** Following multivariate adjustment, the subjects in the highest quartile exhibited a 46% reduction in the risk of CMRFs, a 33% reduction in the risk of diabetes, a 31% reduction in the risk of hypertension, and a 36% reduction in the risk of hyperlipidemia, compared with those in the lowest quartile. Furthermore, each 1-unit increase in OBS was remarkably negatively correlated with the prevalence of CMRFs, diabetes, hypertension, and hyperlipidemia. The correlation between OBS and CMFRs was found to be mediated by serum γ-glutamyltransferase (GGT) and white blood cells (WBC), and the mediation effect of GGT levels and WBC, accounting for 6.90% and 11.51%, respectively. Lastly, the multivariate Cox regression model revealed that elevated OBS, irrespective of whether it was treated as a categorical or continuous variable, exhibited a significant association with decreased mortality from all causes, cardiovascular disease, and cancer.

**Conclusions:** An increased OBS might reflect a lower risk of CMRFs and a favorable prognosis for individuals with CMRFs. Moreover, WBC and GGT may play a potential mediating role between OBS and CMRFs.

**What is new?:**

- An elevated OBS was correlated with a reduced risk of developing CMRFs. Furthermore, WBC and GGT could act as mediators in this association.
- An increased OBS might indicate a positive prognosis for individuals with CMRFs.

**What are the clinical implications?:** OBS might serve as a valuable tool to guide the general population toward adopting healthy diets and positive lifestyle patterns in order to mitigate the risk of CMRFs and enhance the prognosis of CMRFs individuals with CMRFs.

## 1. Introduction

Cardiometabolic risk factors (CMRFs), including hypertension, hyperglycemia, hyperlipidemia, and obesity, are recognized as risk factors for the development of cardiovascular disease (CVD). Indeed, these four CMRFs were collectively responsible for over 60% of global CVD-related mortality in 2010 [1]. Hence, the implementation of early preventive strategies targeting CMRFs can effectively conduce to a reduction in mortality rates.

Although the pathogenesis through which CMRFs facilitate the development of CVD is intricate and multifactorial, a growing body of evidence suggests that oxidative stress serves as a prevalent and fundamental mechanism [2, 3]. Modifiable behavioral risk factors, including smoking, alcohol consumption, an unhealthy diet, and physical inactivity, have been found to be associated with not only CMRFs but also CVD [4, 5]. Smoking and alcohol consumption have been identified as potential oxidants, whereas exercise and a healthy dietary intake (dietary fiber, carotenoids, calcium, etc.) are deemed as potential antioxidants [6]. The oxidative balance score (OBS) is a comprehensive metric that represents the overall oxidative effects of prooxidants and antioxidants in terms of dietary and lifestyle levels. The greater the OBS suggests that the antioxidative components exhibit a greater advantage compared to prooxidant components [7]. Previous findings have established that OBS functions as a biomarker for both oxidative stress and inflammation. These researches have yielded evidence indicating a negative correlation between elevated OBS levels and decreased levels of inflammatory markers such as C-reactive protein (CRP) and white blood cells (WBC) [8], as well as oxidative stress biomarkers, specifically serum γ-glutamyltransferase (GGT) [9]. A study based on Spanish university students implied that a high level of OBS could reduce the risk of all-cause mortality, including CVD-associated and cancer-related death [10]. Besides, prior findings also have reported that increased OBS was correlated with a decreased likelihood of developing diabetes [11, 12] and new-onset hypertension [13]. Notably, CMRFs share a common pathogenic pathway. Thus, evaluating the correlation between any CMFRs and OBS is insufficiently comprehensive. Furthermore, considering that CMFRs account for the majority of CVD-related deaths, exploring the correlation between OBS and mortality among subjects with CMRFs holds significant implications for tertiary preventive strategies.

To address this question, we utilized data derived from the National Health and Nutrition Examination Survey (NHANES). Firstly, a cross-sectional study was designed to explore the correlation between OBS and CMRFs and further evaluate the possible mediating roles of inflammation and oxidative stress indicators in this relationship. Next, a longitudinal study was undertaken to examine the correlation between OBS and mortality among subjects with CMRFs, including both overall and disease-specific outcomes.

## 2. Materials and methods

### 2.1. Study population

The data utilized in this analysis were acquired from NHANES, which employed complex, stratified, and multistage probability sampling techniques to represent the health condition of the general population in the United States. The aforementioned data is readily accessible on the official website of the NHANES (https://www.cdc.gov/nchs/nhanes/).

Part I: The present study included a total of 101316 participants spanning ten consecutive NHANES cycles (1999-2018). Following the exclusion of participants younger than 20 years, pregnant women, and those without eligible OBS and CMRF data, a total of 29289 subjects were ultimately selected (Supplementary Figure 1). Among them, 23190 individuals were diagnosed with CMRFs. An analysis was implemented to evaluate the cross-sectional correlation between OBS and CMRFs.

Part II: We additionally included participants with a confirmed diagnosis of CMRFs, subsequently excluding 28 subjects due to loss of follow-up. Among the 23162 individuals with CMRFs, 3292 individuals experienced all-cause mortality by December 31, 2019 (Supplementary Figure 1). Thereafter, a longitudinal analysis was implemented to investigate the relationship between OBS and mortality.

### 2.2. Definition of CMRFs and mortality

Part I: CMRFs were defined as hypertension, diabetes, hyperlipidemia, or any combination of the three conditions. Diabetes was ascertained through self-reported diagnosis, utilization of diabetes medications, hemoglobin A1c levels equal to or exceeding 6.5%, or fasting plasma glucose levels equal to or exceeding 126 mg/dl. Hypertension was identified by self-reported diagnosis, utilization of anti-hypertensive medications, or systolic/diastolic blood pressure greater than or equal to 140/90 mmHg. Hyperlipidemia was diagnosed through self-reported diagnosis, triglyceride ≥ 150 mg/dL, total cholesterol ≥ 200 mg/dL, high-density lipoprotein cholesterol levels < 40 mg/dL for males or <50 for females, or low-density lipoprotein cholesterol levels ≥ 130 mg/dL.

Part II: Survival status and causes of death were ascertained by accessing the National Death Index file (https://www.cdc.gov/nchs/data-linkage/mortality-public.htm). ICD-10 was employed to ascertain the underlying causes of death among the participants, including mortality due to CVD (heart disease: I00– I09, I11, I13, I20–I51) and malignant neoplasms (C00–C97). The duration of follow-up was calculated by estimating person-years from the time of the interview until either the occurrence of death or December 31, 2019, whichever transpired first.

### 2.3. OBS assessment

The calculation of OBS for each participant was derived from prior reports [14, 15]. In accordance with this methodology, a comprehensive selection of 16 dietary and 4 lifestyle components were identified, all of which exhibit an association with oxidative stress. The specific evaluation methods for OBS are detailed in Supplementary Table 1. The dietary intake of each participant was determined by two 24-hour dietary recall surveys whenever data for two days were available. In cases where only one day of dietary recall data was available, data from the first day was used for calculation purposes. The participants in this study were allocated points ranging from 0 to 2 for dietary antioxidants based on the distribution of tertiles within the study population. Tertile 1 to Tertile 3 were assigned ascending point values. Conversely, a reversed scoring system was utilized for dietary pro-oxidants. Participants in Tertile 1 received the highest score of 2 points, whereas those in Tertile 3 obtained the lowest score of 0 points. Lifestyle component scores were computed using the following methodology: cotinine levels were employed to estimate smoking status and allocate scores, ranging from 0 to 2, to Tertile 3 through Tertile 1, respectively. Body Mass Index (BMI) was computed by dividing the weight (in kilograms) by the square of the height (in square meters) and subsequently stratified into three categories: 2 points (<25 kg/m^2^), 1 point (25-29.9 kg/m^2^), or 0 points (≥30 kg/m^2^). Non-drinkers participants were allocated 2 points, whereas individuals who consumed alcohol within the range of 20 g/day for females and 30 g/day for males were awarded 1 point. Conversely, those who exceeded the aforementioned limits, that is, consuming over 20 g/day for females and over 30 g/day for males, were assigned 0 points [11]. Physical activity was evaluated as weekly metabolic equivalents (METs) and awarded a score of 0 (<7.5 METs-h/wk), 1 (7.5-30 METs-h/wk), and 2 (>30 METs-h/wk) [11].

### 2.4. Measurement of GGT and WBC

The Coulter DxH 800 analyzer was utilized to measure WBC counts, which were subsequently repor*ted* as ×1000 cells/ *µ*L. The enzymatic rate method was employed to analyze GGT levels.

### 2.5. Covariates

The collection of baseline data was accomplished through conducting interviews and laboratory tests, including general information [age, gender, race/ethnicity, marital status, educational attainment, family poverty-to-income ratio (PIR)], dietary factors (intakes of total energy, caffeine, and sodium), and laboratory data [uric acid, estimated glomerular filtration rate (eGFR) and alanine aminotransferase (ALT) level]. The eGFR was determined by employing the Chronic Kidney Disease Epidemiology equation [16].

### 2.6. Statistical analysis

All statistical analyses were conducted in compliance with NHANES analytical guidelines. No variable exhibited a missing data rate exceeding 10%, and multiple imputations were conducted to address missing values [17]. Continuous variables were expressed as the weighted mean ± standard error and analyzed using a one-way ANOVA. Categorical data were presented as weighted percentages, and group comparisons were assessed using the Chi-square test.

We implemented multivariable logistic regression analysis to analyze the relationship between OBS and CMRFs, in addition to utilizing multiple Cox proportional hazards regression analysis to investigate the correlation between OBS and mortality due to all causes, CVD, and cancer. Three distinct models were constructed, with age, sex, and race/ethnicity adjusted in Model I; the remaining demographic information was incorporated in Model II; intakes of total energy, caffeine, sodium, uric acid, eGFR, and ALT were introduced in Model III. The OBS variable was constructed as a continuous variable and quartiles (Q1-Q4), with the lowest quartile serving as the reference category. Additionally, potential correlations between dietary/lifestyle OBS, CMRFs, and mortality were analyzed. Restricted cubic spline (RCS) with 4 knots was utilized to visualize the dose-response relationship between OBS and the risk of CMRFs and mortality.

We employed mediation analysis to assess the potential mediating influence of GGT levels and WBC counts on the relationship between OBS and CMRFs. The mediation analysis was conducted utilizing R package mediation, and confounding variables, including general information, dietary factors, and laboratory data, were adjusted for.

To evaluate the robustness of the results, a series of sensitivity analyses were implemented. To begin, given the relationship between different dietary patterns and CMRFs, the Dietary Approaches to Stop Hypertension (DASH) and Healthy Eating Index-2015 (HEI-2015) scores were introduced into the fully adjusted model. Secondly, subjects who experienced mortality within a two-year frame were excluded to mitigate the potential impact of reverse causality. Ultimately, we censored the follow-up at 10 years. We implemented all statistical analyses using R software (version 4.2.2). *P* values < 0.05 were considered statistically significant.

## 3. Results

### 3.1. Correlations between OBS and CMRFs

A total of 29,289 eligible participants were chosen for this analysis, aiming to examine the association between OBS and CMRFs. Among these participants, 4,233 individuals were diagnosed with diabetes, 11,736 individuals were diagnosed with hypertension, and 20,823 subjects were diagnosed with hyperlipidemia. Table 1 summarizes the main characteristics of participants based on OBS quartiles. The results indicated a higher levels of OBS and a trend for a lower prevalence of CMRFs, diabetes, hypertension, and hyperlipidemia (all *P*<0.001). As OBS increased, subjects were more likely to be male, non-Hispanic white, married, possess higher education levels, have a higher PIR, elevated levels of uric acid, and a higher intake of sodium and total energy. Notably, individuals with elevated levels of OBS exhibited increased ALT levels and caffeine consumption. Besides, differences were noted in age and eGFR levels among the groups. Supplementary Table 2 presents the individual components of OBS according to the OBS quartile. As OBS increased, there was a corresponding progressive increase in the consumption of dietary fiber, carotene, vitamin B12, vitamin C, vitamin E, vitamin B6, calcium, magnesium, zinc, copper, selenium, riboflavin, niacin, total folic acid, total fat, and iron. Likewise, there was a gradual increase in the duration of physical activity, whereas BMI and cotinine levels exhibited a gradual decrease (all *P*<0.001).

Table 2 lists the outcomes of the three multivariable logistic regression models employed to investigate the relationship between OBS, evaluated as both a continuous and quartile variable, and the risk of CMRFs. After multivariable adjustment, participants in Q2, Q3, and Q4 exhibited a risk reduction of 21%, 36%, and 46%, respectively, for CMFRs, compared to those in the lowest quartile (*P* for trend < 0.001). After adjusting for all covariates, elevated OBS was found to be significantly correlated with a decreased risk of diabetes, hypertension, and hyperlipidemia (all *P* for trend <0.001). More specifically, there was a 33% reduction in the risk of diabetes, a 31% reduction in the risk of hypertension, and a 36% reduction in the risk of hyperlipidemia in Q4 relative to Q1. The multivariable model presented a negative association between a one-unit increment in OBS and the prevalence of CMRFs, diabetes, hypertension, and hyperlipidemia (all *P*<0.001). Meanwhile, RCS demonstrated a significant negative linear association between OBS and CMRFs (*P* for nonlinear=0.120, Figure 1A). As portrayed in Table 3, after adjusting for all confounders, each 1-unit increase in dietary/lifestyle OBS was significantly and inversely linked to the risk of CMRFs, diabetes, hypertension, and hyperlipidemia (all *P*<0.001).

Noteworthily, sensitivity analysis exposed no significant change in the results, even after further adjusting for DASH and HEI-2015 scores (Supplementary Table 3). Notably, the mediation analyses unveiled significant mediating effects of GGT and WBC in the relationship between OBS and CMRFs, accounting for 11.51% and 6.90% of the total mediation proportion, respectively (*P*<0.01, Table 4).

### 3.2. The association of OBS with mortality among subjects with CMRFs

Out of a total of 23,162 subjects with CMRFs in this study, 3,292 individuals succumbed to all-cause mortality, 835 individuals died due to CVD, and 789 individuals passed away as a result of cancer. RCS indicated that the correlation between OBS and all-cause mortality conformed to a dose-response pattern (*P* for nonlinear=0.153, Figure 1B). Table 5 presents the results of the three different Cox regression models investigating the correlation between OBS and all-cause, CVD-related, and cancer-associated deaths. In the fully adjusted model, the linear correlation between OBS and all-cause mortality continued to exhibit significance. The multivariable-adjusted hazard ratios and corresponding 95% confidence intervals for Q2, Q3, and Q4 were 0.89 (0.79, 1.00), 0.85 (0.75, 0.96), and 0.65 (0.57, 0.74) respectively, compared to Q1 (P for trend <0.001). After full adjustment, individuals in Q4 experienced a significant decrease in the risk of CVD mortality by 40% and cancer mortality by 42% in comparison to those in Q1. Meanwhile, analyzing OBS as a continuous variable exposed that every unit increase in OBS was inversely related to all-cause, CVD-related, and cancer-related deaths (all *P*<0.05). Table 6 illustrates similar results in the correlation analysis between dietary/lifestyle OBS and mortality. Lastly, the multivariable model suggested that each incremental unit increase in dietary/lifestyle OBS was linked to a reduced risk of all-cause, CVD-related, and cancer-related mortality (all *P*<0.05).

Regarding sensitivity analyses, the addition of DASH and HEI-2015 scores in the multivariate analysis did not substantially affect the results. Nonetheless, the correlation between OBS and CVD mortality was decreased after adjusting for HEI-2015 scores (Supplementary Table 4). Besides, there was no significant change in the correlation between OBS and all-cause, CVD-related, and cancer-related deaths, even after the exclusion of individuals who died within the two-year frame or after censoring the data at a ten-year follow-up period (Supplementary Table 4).

## 4. Discussion

This study employed both cross-sectional and longitudinal designs and discovered a negative linear relationship between OBS and CMRFs in United states (US) adults, irrespective of whether the former was modeled as a categorical or continuous variable. Furthermore, mediation analyses indicated significant mediating effects of GGT and WBC in this relationship. Moreover, the cohort study showed that high OBS was linked to a decreased risk of all-causes, CVD-induced, and cancer-related deaths among individuals with CMFRs.

Hypertension, diabetes, and hyperlipidemia interact with each other and share a common pathogenic pathway. Our study is the first to link these three diseases and explore the correlation between CMFRs and OBS. Our results presented that increased OBS was related to a decreased risk of CMFRs. Previous researches have established an negative correlation between OBS and low-density lipoprotein cholesterol levels, diabetes, and new-onset hypertension [8, 11, 13]. A cross-sectional study implemented in South Korea revealed a significant negative correlation between OBS and the risk of developing metabolic syndrome (MetS) [18]. Conversely, a subsequent study performed in Iran found no significant relationship between OBS and MetS in adults [19]. Our results extended these prior findings, corroborating that high OBS is not only remarkably linked to a decreased risk of hypertension, diabetes, and hyperlipidemia but also significantly correlated with a lower risk of CMRFs in US adults. Furthermore, our study revealed an inverse relationship between both dietary OBS and lifestyle OBS and the prevalence of hypertension, diabetes, hyperlipidemia, and CMRFs. Multiple findings have explored the effect of dietary antioxidants on these three CMRFs, determining that individuals with elevated levels of dietary antioxidant capacity exhibit a reduced likelihood of developing hypertension, diabetes, and dyslipidemia in comparison to those with decreased levels of dietary antioxidant capacity [20-23]. Furthermore, a study based on the Chinese population has demonstrated a negative relationship between lifestyle OBS and CMFRs [24]. Consequently, adhering to a healthy lifestyle has the potential to mitigate the risk of developing diabetes, hypertension, and hyperlipidemia [25-27]. Hence, we postulate that OBS can serve as a valuable tool to guide the general population toward adopting healthy diets and positive lifestyle patterns in order to reduce the occurrence of CMRFs.

Consistent with the observations of prior studies [11, 15, 28], dietary prooxidants (iron and total fat) exhibited an upward trend with a rise in OBS, which was in contradiction with the OBS assignment scheme. This discrepancy might be ascribed to higher total energy intake. While a single antioxidant or prooxidant may be involved in the etiology and development of CMRFs, it is crucial to consider their antagonistic or synergistic effects. Therefore, OBS was used as a comprehensive assessment tool to evaluate oxidative balance in individuals and explore its impact on CMRFs. We observed a dose-response relationship between OBS and the risk and prognosis of CMRFs. Consistent with our results, a cohort study reported a significant inverse dose-response correlation between elevated OBS and the occurrence of new-onset hypertension [13]. Similarly, another study reported a progressive decline in the risk of all-cause mortality as the intake of dietary OBS increased [29]. In summary, our findings provide compelling evidence to endorse a public health recommendation that dietary and lifestyle antioxidants can effectively mitigate the risk of CMRFs and enhance the prognosis of patients. Although the precise mechanism by which OBS impacts CMRFs remains elusive, oxidative stress assumes a significant role. Oxidative stress refers to the disruption of the equilibrium between intracellular antioxidants and prooxidants, frequently accompanied by the accumulation of reactive oxygen species (ROS) [30], which in turn drives insulin resistance and lipid peroxidation through multiple mechanisms, thereby exerting adverse effects on blood pressure, glucose, and lipid metabolism and increasing the risk of CVD [30-34]. What’s more, ROS contributes to the development of hypertension by inducing endothelial damage, vascular dysfunction, and remodeling, as well as sympathetic nervous system excitation [35]. In addition, the occurrence of oxidative stress triggers a reduction in nitric oxide levels (NO), eventually resulting in endothelial dysfunction and subsequently increasing the risk of CVD and hypertension [36]. Indeed, NO plays a pivotal role in carbohydrate metabolism, and a decrease in its bioavailability can further aggravate insulin resistance [37]. In conjunction with oxidative stress, activation of the inflammatory response also plays a decisive role in the pathophysiology of CMRFs. Inflammatory mediators stimulate immune cells activation within target organs, thereby exacerbating vascular dysfunction, remodeling, and fibrosis, collectively leading to elevated blood pressure [38]. Chronic low-grade inflammation is causally correlated with insulin resistance, as well as the activation and infiltration of immune cells, thereby promoting the occurrence and development of diabetes [39, 40]. Moreover, CMFRs increase ROS production, limit NO generation and trigger inflammatory responses [41, 42]. The interaction between oxidative stress, inflammatory response, and CMRFs is multidirectional and complex. Overall, it is reasonable to speculate that inflammatory response and oxidative stress may act as potential mediators between OBS and CMRFs. Our research substantiates the aforementioned viewpoint, revealing that the relationship between OBS and CMRFs is partially mediated by GGT and WBC.

As is well documented, individuals with CMRFs have a poor prognosis. In this study, high OBS was found to be related to a decreased risk of all-cause, CVD-related, and cancer-related mortality among individuals with CMRFs. The results of our study align with a previous Spanish study, which described a negative correlation between OBS and mortality [10]. A prospective study conducted among adults aged 45 years and above in the US found showed that individuals with high OBS might potentially experience a lower all-cause mortality risk [43]. In addition, we further validated that individuals who are exposed to higher levels of dietary or lifestyle OBS may potentially exhibit a reduced risk of mortality. Consistent with our study results, Wang et al. demonstrated that dietary OBS was negatively linked to deaths from all causes and CVD [29]. According to a meta-analysis, adhering to a healthy lifestyle has been found to significantly decrease the mortality rate among middle-aged women [44]. Taken together, the results of the current study possess significant implications for public health, particularly in terms of devising strategies to mitigate the risk of CMRFs and enhance the prognosis of CMRFs individuals with CMRFs.

Despite the support from substantial sample size and a long-term NHANES follow-up, certain limitations in our research should be acknowledged. To begin, the cross-sectional nature of our study inherently limited our ability to establish causal relationships between OBS and CMFRs. Secondly, self-reported questionnaires may lead to biases in assessing dietary OBS. Thirdly, our analysis exclusively considered baseline OBS without accounting for the potential diet and lifestyle modifications of participants during the follow-up process. Regrettably, the limitations of NHANES precluded dynamic adjustments in OBS. Finally, despite conducting a multivariate analysis, the possibility of selection bias influencing our findings cannot be excluded.

## 5. Conclusion

This cross-sectional and cohort study demonstrated that an increased OBS has the potential to decrease the risk of CMRFs and enhance the prognosis of individuals with CMRFs. Additionally, our study revealed that the GGT and WBC may potentially act as mediators in the association between OBS and CMRFs.

## Data Availability

All data can be obtained on the NHANES official website.

## Ethical standards

The NHANES survey was granted approval by the institutional review board of the National Center of Health Statistics, and all respondents provided written informed consent prior to their participation.

